# Genomic epidemiology of the Los Angeles COVID-19 outbreak

**DOI:** 10.1101/2020.09.15.20194712

**Authors:** Longhua Guo, James Boocock, Evann E. Hilt, Sukantha Chandrasekaran, Yi Zhang, Chetan Munugala, Laila Sathe, Noah Alexander, Valerie A. Arboleda, Jonathan Flint, Eleazar Eskin, Chongyuan Luo, Shangxin Yang, Omai B. Garner, Yi Yin, Joshua S. Bloom, Leonid Kruglyak

## Abstract

Los Angeles (LA) County has sustained a large outbreak of severe acute respiratory syndrome coronavirus 2 (SARS-CoV-2). To learn about the transmission history of SARS-CoV-2 in LA County, we sequenced 142 viral genomes from unique patients seeking care at UCLA Health System. 86 of these genomes are from samples collected before April 19, 2020. We found that the early outbreak in LA, as in other international air travel hubs, was seeded by multiple introductions of strains from Asia and Europe. We identified a US-specific strain, B.1.43, which has been found predominantly in California and Washington State. While samples from LA County carry the ancestral B.1.43 genome, viral genomes from neighbouring counties in California and from counties in Washington State carry additional mutations, suggesting a potential origin of B.1.43 in Southern California. We quantified the transmission rate of SARS-CoV-2 over time, and found evidence that the public health measures put in place in LA County to control the virus were effective at preventing transmission, but may have been undermined by the many introductions of SARS-CoV-2 into the region. Our work demonstrates that genome sequencing can be a powerful tool for investigating outbreaks and informing the public health response. Our results reinforce the critical need for the U.S. to have coordinated inter-state responses to the pandemic.

## Introduction

Since the first report of pneumonia patients associated with a novel coronavirus in Wuhan, China in late December, 2019 (Zhu et al., 2020), SARS-CoV-2 has spread across the globe, infecting 29 million people, and killing 927 thousand as of September 14th, 2020. The United States (US) alone has reported 194 thousand deaths (Dong et al., 2020). Genomic surveillance via viral genome sequencing is crucial for determining outbreak dynamics, detecting viral evolution and informing public health interventions. Studies in Washington State showed that most infections there likely stemmed from a single introduction event of strain WA1, followed by cryptic community transmission (Bedford et al., 2020), while sequencing of viral genomes from Northern California and New York City demonstrated that there had been multiple independent introductions into these areas from international and domestic travelers (Deng et al., 2020)(Gonzalez-Reiche et al., 2020). Viral genomes from samples collected during the period from March 22 to April 15 at the Cedars Sinai Medical Center also showed multiple introductions of SARS-CoV-2 into Los Angeles County (Zhang et al., 2020).

The first complete genomes of SARS-CoV-2 from the root of the pandemic in Wuhan, China were deposited into GISAID and GenBank in January 2020 (Zhu et al., 2020)(Wu et al., 2020) (Zhou et al., 2020). Since then, large-scale global efforts to sequence SARS-CoV-2 have led to 62,6441 genomes in GISAID as of July 14, 2020. Most of these genomes were obtained using meta-transcriptomic sequencing or nanopore sequencing (Quick et al., 2017) (Elbe and Buckland-Merrett, 2017a) (Shu and McCauley, 2017a). These approaches are costly and resource-intensive for individual laboratories.

We recently developed a rapid and inexpensive sequencing method based on targeted reverse transcription of the SARS-CoV-2 genome directly from patient RNA (Guo et al., 2020). We used this method, together with a meta-transcriptomic approach, to generate 142 high-quality viral genome sequences from patients residing in LA County who were seen at UCLA Health. We used these genomes, together with publicly available ones, to investigate the history of the SARS-CoV-2 outbreak in LA County.

## Results

### Rapid low-cost sequencing of SARS-CoV-2 genomes

We recently developed V-seq, a sequencing method that uses virus-specific RT primers tiled across the SARS-CoV-2 genome for viral sequence enrichment (Guo et al., 2020). The V-seq protocol is more rapid and 10 times cheaper than commercially available meta-transcriptomics approaches (*e.g*., NEBNext Ultra II). We sequenced 122 patient samples from UCLA Health with V-seq and 138 samples with NEBNext Ultra II (Figure S1, Table S1). We obtained 97 and 63 high-quality genomes by V-seq and NEB, respectively (Figure S2, Table 1). For both methods, samples with a higher amount of viral RNA, as determined by the cycling threshold (Ct) of the RT-qPCR used to detect the presence of the virus, had a higher fraction of reads aligning to SARS-CoV-2. To assess the accuracy of V-seq for variant identification, we compared high-confidence variant calls in 18 samples from which we recovered high-quality genomes with both methods. We did not find any discrepancies among 6,657 high-confidence genotype calls at 380 sites across the SARS-CoV-2 genome (Figure S3). These results show that V-seq is a highly accurate approach for sequencing SARS-CoV-2 genomes.

**Table 1:** Summary of the UCLA Health SARS-CoV-2 genomes dataset.

|  | **Average**<br>**Coverage ≥**<br>**3x per library** | **Median**<br>**number of**<br>**reads per**<br>**library** | **Number of**<br>**libraries** | **Passing QC** |
| --- | --- | --- | --- | --- |
| NEB | 0.629 | 3298304 | 138 | 63 |
| V-seq | 0.853 | 3663474 | 122 | 97 |

### Multiple introductions of SARS-Cov-2 into LA County

We obtained 142 new SARS-CoV-2 genomes from samples collected in LA County between February 28 and June 22, 2020 (Figure S4). We combined these genomes with another 144 genomes from LA County obtained from GISAID on July 14, 2020. We performed a phylogenetic analysis with NextStrain using these 286 LA County genomes together with 3,809 genomes sampled from across the world (Hadfield et al., 2018a). LA County genomes were distributed throughout the resulting phylogenetic tree, consistent with multiple independent introductions (Figures 1A, S5A). We used a parsimony-based approach to identify 145 distinct introductions of SARS-CoV-2 into LA county (Figure 1B, S5B). One introduction is related to a large community outbreak cluster containing 58 LA County genomes, which we assigned to the US-specific lineage B.1.43. 33 introductions are related to clusters with more than one LA County genome, and the remaining 111 introductions are found in clusters containing only a single LA County genome, with no evidence of community transmission in our sample.

**Figure 1:**
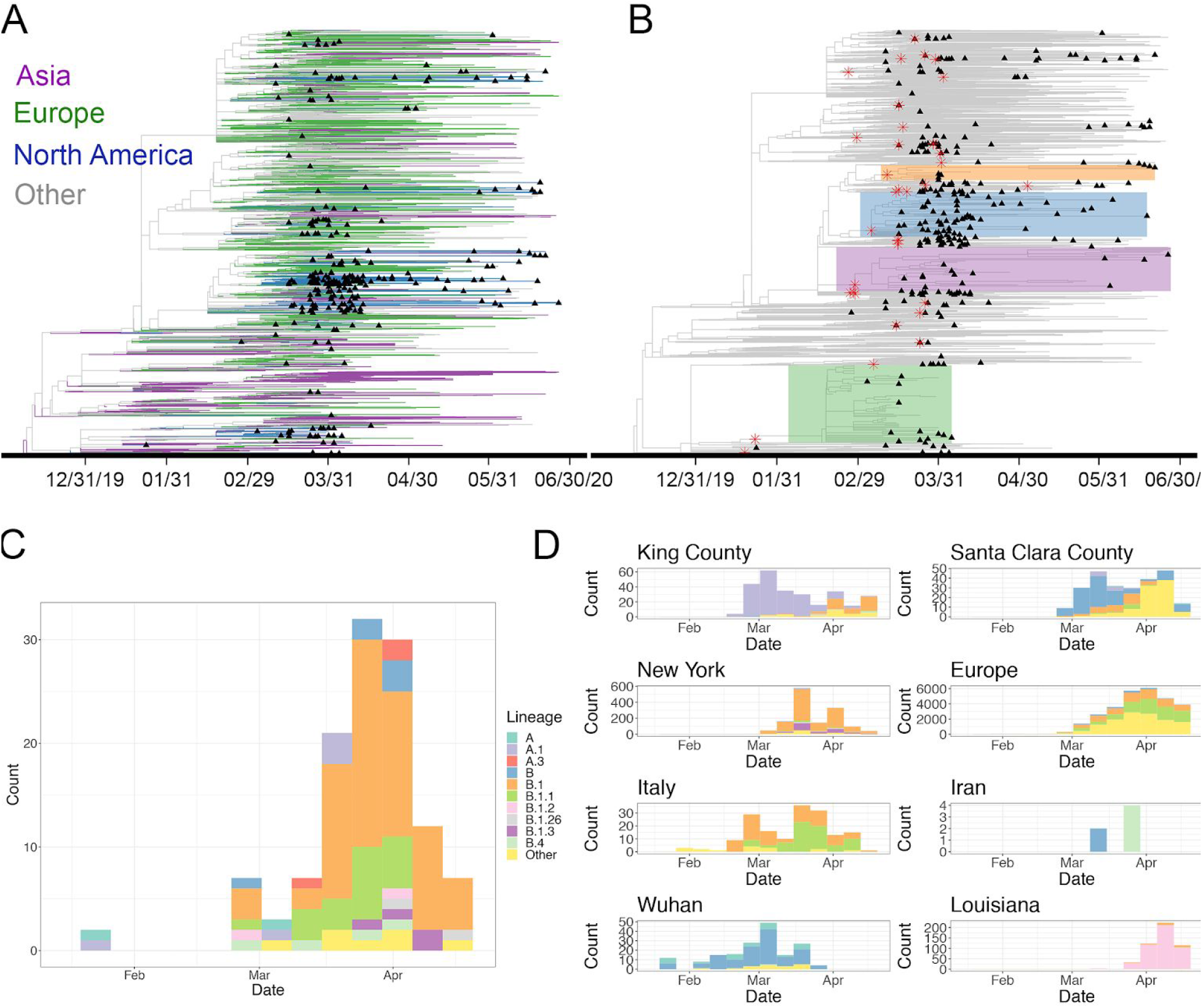
Multiple introductions of SARS-CoV-2 into LA county. **A)** Phylogenetic tree of 286 LA county SARS-CoV-2 genomes together with 3,809 genomes sampled from across the world. Branches are colored according to the region of origin. Tip triangles (black) indicate the position of LA county sequences. **B)** The same tree as in **A** with different annotations. We zoomed in and highlighted four regions (blue, orange, purple, and green) of the tree where a single introduction was related to a cluster containing LA County genomes indicating community transmission. Nodes shown with a red star indicate a LA County introduction related to a cluster with more than one LA County genome. **C)** Assignment of LA County introductions before April 19th, 2020 to lineages. **D)** Observed frequency of different lineages from eight other COVID-19 hotspots from throughout the US and the world.

We estimated that 122 introduction events occured before April 19, 2020. We assigned these introductions to 17 distinct lineages related to global and early U.S. outbreaks (Figure 1C-D, Table S2-3) (Oster et al., 2020). Ninety nine (81%) of these early introductions were assigned to European-derived lineages, whereas the remaining introductions were assigned to Chinese-derived lineages. The earliest introduction of SARS-CoV-2 in LA County involved the A lineage, which represents the root of the pandemic in Wuhan, China. Next, the derived A.1 lineage was introduced. This lineage was common in Washington State during the early outbreak in King County. At around the same time, the B lineage and its derivatives were introduced. We observed that different B lineages were introduced into LA County at around the time of the outbreak of each lineage in a geographic hotspot (Oster et al., 2020). For example, B.1 was introduced into LA County during its outbreak in Italy and New York, while B.1.2 was introduced into LA County during its outbreak in Louisiana. These observations suggest that SARS-CoV-2 has been repeatedly introduced into LA County by a diverse mix of domestic and international travelers.

### The history of a US specific SARS-CoV-2 lineage

Worldwide, a total of 247 B.1.43 samples, including our LA County genomes, have been reported in GISAID as of July 14, 2020. Three were found outside the U.S. as early as March 17. Of the 244 U.S. B.1.43 samples, 67% were found in California and 28% were found in Washington State (Figure 2A, Table S4). The remaining 5% were found in 8 states, including those that neighbour California or each other (Arizona, New Mexico, Utah and Texas) and those on the East Coast (New York, Maryland, Kentucky and Wisconsin). Of the 163 cases in California, 77% were found in LA and neighbouring counties in Southern California, with the largest number of cases in LA County (N=48) (Figure 2B). Of the 67 samples in Washington State, 82% were found in three neighbouring counties in Northern Washington, with the largest number of cases in Whatcom county (N=43) (Figure 2C). The other cases in Washington State do not have associated county information.

**Figure 2:**
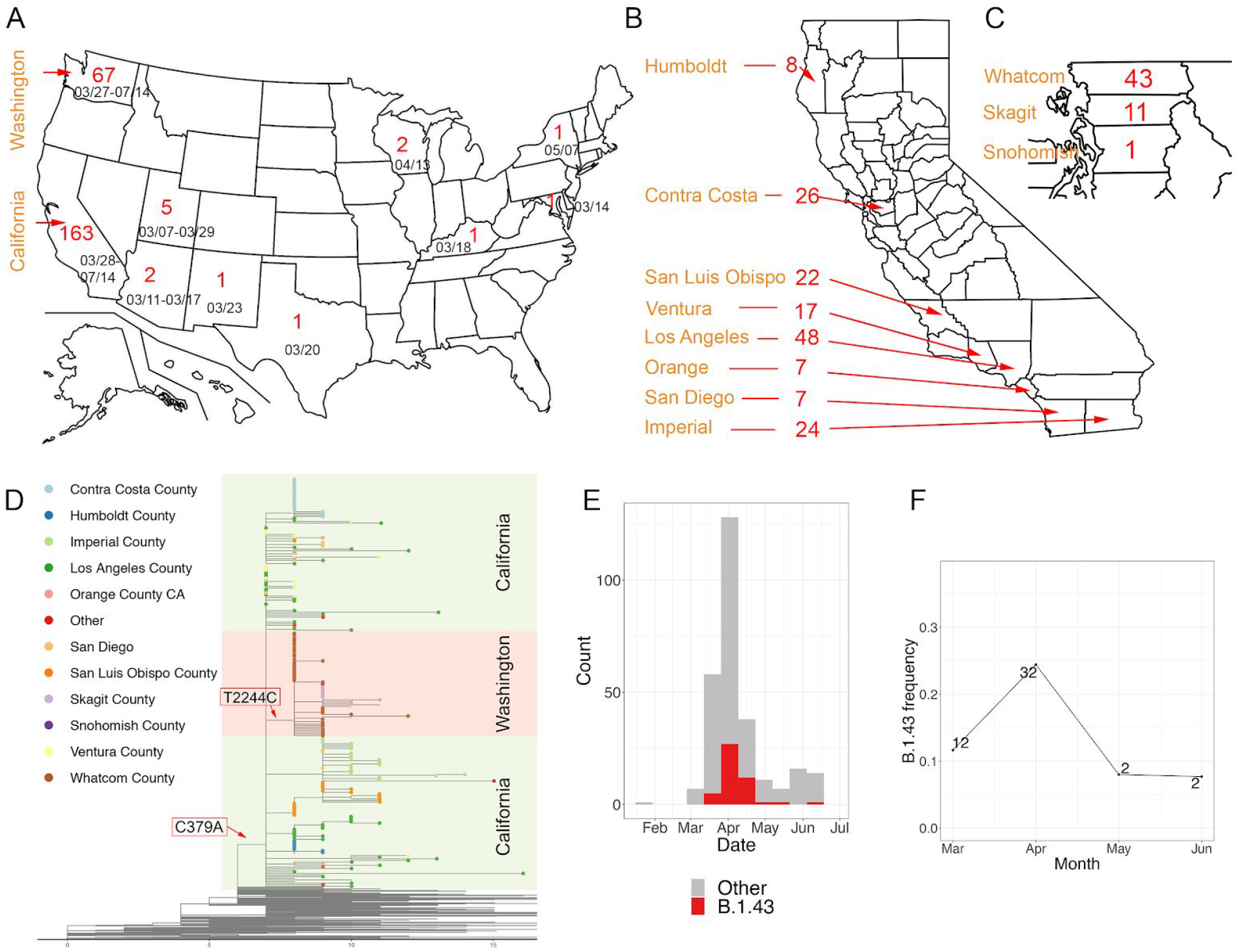
The history of the US-specific lineage B.1.43. **A)** US map showing the states where the lineage B.1.43 has been found. Numbers are reported cases of B.1.43. CA: California; WA: Washington. **B-C)** Map of California (B) and Washington State (C). Counties and the number of B.1.43 cases were labeled. **D)** The B.1.43 strains found in Washington state (orange) are derived from the ancestral B.1.43 sequence by one mutation (T2244C). Some of the B.1.43 strains LA County (dark green) carried the ancestral haplotype. X-axis is the number of mutations compared to the root in Wuhan, China. The nodes are colored based on the US counties from which they were collected. **E)** The distribution of LA County genomes over time. **F)** The frequency of B.1.43 in LA County summarized per month. The number of B.1.43 strains identified each month is written near the data point.

To gain insight into the origin of B.1.43 in the US, we performed a phylogenetic analysis of the 247 B.1.43 and 987 other SARS-CoV-2 genomes sampled from around the world. B.1.43 lineages differ from B.1 at a single position, C379A (ORF1a, L64L) (Figures 2D, S6). All 67 B.1.43 strains from Washington State had at least one additional derived mutation, with 65 sharing the same derived mutation, T2244C (ORF1a, V660A). Genomes with the ancestral B.1.43 sequence were identified only in LA County, three nearby counties (Ventura, San Diego, and Orange), Utah, Kentucky, and Australia. The earliest ancestral B.1.43 sequences were found in Utah on March 7, 2020 and in Southern California on March 28, 2020. The mutational signature and the geographical distribution of B.1.43 strains suggest that California or Utah may have been the source of the Washington State B.1.43 outbreak.

Longitudinal sampling of viral genomes in LA County from February to June allowed us to track the history of the B.1.43 lineage over time. We inferred that the B.1.43 lineage was introduced into LA County once, around March 7, 2020 (95% CI=March 4th, 2020 - March 9th, 2020), and that following its introduction, its frequency among the circulating strains has changed, peaking at ~25% in April and dropping to ~8% in May and June (Figure 2E-F). This result suggests that the B.1.43 lineage is being replaced by other lineages introduced more recently.

### Genomic assessment of the effectiveness of local public health measures

To assess the effectiveness of public health measures put in place in LA County, we used a Bayesian approach (Stadler et al., 2013) to quantify changes in the rate of transmission of the B.1.43 lineage overtime in LA County. The estimate of the effective reproductive number (Re) of this lineage rose to ~5.94 (95% CI=3.1-9.3; Methods) in early March, but was near 1 (95% CI=0.14-2.8) by the middle of April (Figure 3). These results suggest that the “Safer at Home” order put into place in LA County on March 20, 2020, together with other social distancing measures, were effective at reducing the transmission of the virus.

**Figure 3:**
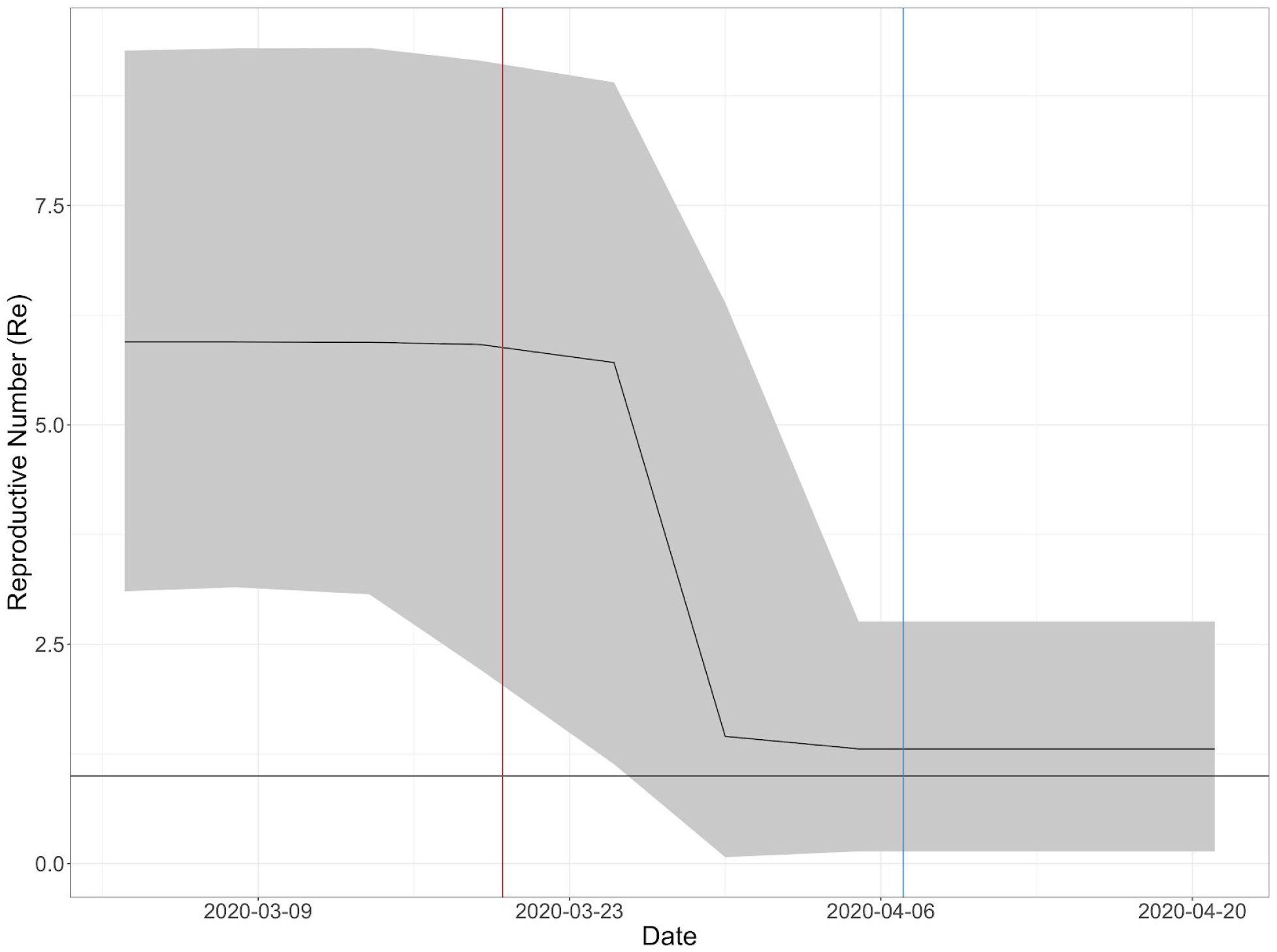
Genomic assessment of public health measures put in place to prevent the spread of SARS-CoV-2 in LA County. Effective reproductive number (Re) quantified across time using all B.1.43 strains from LA County. These genomes were inferred to have arisen from a single introduction event. The grey ribbon shows the 95% highest posterior density (HPD) credible interval for our estimates of Re. The vertical lines depict the time when some public health measures were put in place in LA County. The ‘Safer at Home’ order issued for LA County put in place on March 20, 2020 (red). LA City required masks to be worn when visiting essential businesses on April 7th, 2020 (blue).

## Discussion

We developed a faster and cheaper virus-targeted sequencing approach and applied it to genome sequencing of SARS-CoV-2 samples from a major travel hub, LA County. We combined our 142 genomes with publicly available data and found that SAR-CoV-2 was introduced into LA County many times, likely via a variety of domestic and international travel routes. We studied the history of a US-specific SARS-CoV-2 lineage, B.1.43 by combining mutational signatures and regional distributions, and found evidence that B.1.43 originated in Southern California or Utah and spread to northern Washington State.

A limitation of our study is that we partially relied on publicly available genome sequences for our inferences. Publicly available genomes are sampled at different rates throughout the US and the world. As an example, the lack of B.1.43 lineages outside Washington State and California could reflect a lack of sequencing data from other states. In agreement with another study of LA County genomes (Zhang et al., 2020), we found evidence that SARS-CoV-2 has been introduced many times. However, without detailed travel information, we cannot pinpoint the sources of these introductions or rule out community transmission post-introduction. Finally, the UCLA patient population is affluent relative to all of LA County, and likely to travel more frequently, which suggests that our analysis may overestimate the relative importance of introductions to the overall dynamics of the SARS-CoV-2 outbreak in LA County.

Early in the pandemic, LA County officials followed the advice of public health experts. Schools, bars, and gyms were closed on March 16, 2020, and all non-essential business activity was stopped on March 20, 2020. However, even after these orders were put in place, the number of reported daily cases continued to increase, with an average of ~850 cases per day in April and May (LA Times’ independent count; https://github.com/datadesk/california-coronavirus-data). We analyzed the rate of transmission of SARS-CoV-2 in LA County using the genome sequences, and found evidence that the public health measures were effective in reducing the transmission of the virus. SARS-CoV-2 was repeatedly introduced into LA County from hotspot regions throughout the U.S. and the world (Oster et al., 2020). These ongoing introductions may have undermined the effectiveness of the control measures put in place in LA County (Fauver et al., 2020). Our results reinforce the critical need for the U.S. to coordinate local responses to the SARS-CoV-2 pandemic.

## Data Availability

Analysis code and processing scripts, and snakemake (v5.17.0) pipelines for our analysis are available at https://github.com/theboocock/COVID-NGS2. Virus genome sequences were uploaded to GISAID.

## Acknowledgements

We thank all authors and contributors who have submitted their genome sequences to GISAID. LG performed the experiments. YY developed V-seq with help from LG. LG, JB, and JSB analyzed the data. LG, JB, JSB, and LK wrote the manuscript. All co-authors approved the manuscript.

We thank the UCLA David Geffen School of Medicine’s Dean’s Office for their support, the Fast Grants. Inc for funding of this work. A generous donation was provided by Jane Semel. This work was supported by funding from Howard Hughes Medical Institute (to LK), and Damon Runyon Cancer Research Foundation (DFS-43-20 to YY).

## Materials and Methods

### Sample collection and processing

The clinical samples were submitted to be tested for SARS-CoV-2 at the Virology laboratory at UCLA between Feb 21, 2020 and June 28th, 2020. The samples were tested on one of three diagnostic testing protocols approved for Emergency Use Authorization (EUA) by the Food and Drug Administration (FDA). The three protocols were: CDC 2019-Novel Coronavirus (2019-nCoV) Real-Time RT-PCR Diagnostic Panel, DiaSorin Molecular Simplexa™ COVID-19 Direct or TaqPath COVID-19 Combo Kit. The extracted RNA from these samples were approved to be sequenced by UCLA’s Institutional Review Board (IRB) under studies IRB#20-000527 and IRB#20-001157.

### Raw read processing and alignment

All libraries were sequenced on an Illumina NextSeq 500 Sequencing System. We used bcl2fastq (v2.20.0.422) to obtain libraries for each sample allowing a 1 barcode mismatch for NEB Ultra II samples, and 0 a barcode mismatch for V-seq libraries.

For the V-seq libraries, we removed all custom RT-primers using a custom script written in the R (v4.0.0) programming language (R Core Team, 2020). This script uses the ShortRead (v1.46)(Morgan et al., 2009) package from Bioconductor (Huber et al., 2015). We mapped the reads from each library to a composite reference genome consisting of human (hg38) and SARS-CoV-2 (NC_045512) using the bwa (v0.7.17-r1188) mem command (Li, 2013). For the V-seq libraries from primer sets oP1, oP3 or oP4, we combined the 3 base-pair unique molecular identifier (UMI) with the 6 base random or “not-so-random” hexamer sequence to create a 9 base-pair UMI. For reads assigned to primer set oP2, we combined the 8 base-pair UMI with the 6 base-pair random hexamer to create a 14 base-pair UMI. For a more detailed description of the primer design see (Guo et al., 2020). We used the GroupReadsByUmi tool from the fgbio (v1.2.0; https://github.com/fulcrum-genomics/fgbio) toolkit to group reads using this UMI. We generated molecular consensus sequences using the fgbio CallMolecularConsensusReads tool. For the NEB libraries, PCR duplicates were removed using MarkDuplicates from the Picard tool suite (v2.22.2; (http://broadinstitute.github.io/picard). We calculated the number of reads that mapped to human rRNA, other regions of the human genome, and SAR2-CoV-2 before and after deduplication. We visualized the relationship of these metrics to the cycling threshold (Ct) of the RT-qPCR used to detect the presence of SARS-CoV-2 in each patient sample using ggplot2 (v3.3)(Wickham, 2016).

### Variant calling and consensus sequence generation

We merged reads across unique patient sample and library type combinations and called bases at all sites in each of these samples using the mpileup and call commands of bcftools (1.10.2)(Li et al., 2009). We removed any sites with depth less than 3 or a variant quality (QUAL) of less than 20. We flagged any site called heterozygous in at-least one sample, and calculated the allelic ratio of the alternative allele to the total depth in each sample for these variants. If the allelic ratio was between > 0.1 and < 0.9 and had at least two unique reads supporting each allele, we flagged the variant as being a possible intra-patient variable site. We removed any samples with greater than 4 called intra-patient variable sites. We used bcftools to create consensus sequences and masked any bases that were not found in the filtered VCF file. We also masked any heterozygous sites. Consensus sequences with greater than 80% coverage at a depth of > 3 were considered to have passed quality control.

### Phylogenetic analysis

The available SARS2-CoV-2 genomes were downloaded from GISAID (Accessed July 13th, 2020) (Elbe and Buckland-Merrett, 2017b; Shu and McCauley, 2017b). We filtered these genomes using the Nextstrain (pipeline (Hadfield et al., 2018b). We required these genomes to be at least 25,000 bases in length and have at most 4,500 bases of missing data. We also removed a sequence (USA/CA-ALSR-0513-SAN/2020) which had an incorrect date recorded in GISAID. These filtering steps left us with 59,830 genomes. We assigned lineages to the UCLA Health and publicly available genomes according to a recently proposed nomenclature with Pangolin (https://github.com/cov-lineages/pangolin) (Rambaut et al., 2020).

We performed phylogenetic analysis of all LA County genomes using Nextstrain (v1.16.7) (Hadfield et al., 2018b). In more detail, we combined all LA County genomes with a sampling of genomes from around the world. To achieve this, we utilized proximity sampling and allowed 20 samples per country, year, and month combination, and 10 contextual samples per country and year combination (see https://nextstrain.github.io/ncov/ for a more detailed description of how sampling works in Nextstrain). These genomes were run through the entire Nextstrain pipeline, and we explored the results and exported the trees from the Auspice web application (v2.16.0).

For our focused analysis of the B.1.43 lineage, we combined all genomes assigned to this lineage with a random sampling of genomes from around the world. As before, we utilized proximity sampling but only allowed 2 samples per country per and year combination. We also sampled contextually 2 samples per country and year combination.

To identify the SARS-CoV-2 introduction events in Los Angeles County, we utilized maximum parsimony as implemented in the Castor (v1.6.2) package to infer the value of a two-state character (LA County vs. non-LA County) for every node in the tree (Louca and Doebeli, 2018). When the child of a node was assigned to LA County but the parent was non-LA County, we determined that an introduction event must have happened. We set all ambiguous assignments to non-LA County. We set all polytomies (nodes with greater than two genomes) to non-LA County and any children of this node assigned to LA County were determined to be independent introductions. We assigned lineages to introductions by taking the most common lineage found in the offspring of these nodes.

### Phylodynamics analysis

To investigate how the transmission of SARS-CoV-2 changed overtime in LA County, we used a Bayesian birth-death skyline model implemented in BEAST (v2.5)(Stadler et al., 2013). The HKY + Γ model of nucleotide substitutions was used with a strict molecular clock. The clock rate had a Γ prior distribution with a mean of 8 · 10^−4^ subs/site/year and a standard deviation of 5 · 10^−4^ reflecting estimates for the mutation rate of SARS-CoV-2 (Andersen et al., 2020). We assumed that the infectious period was 10 days, which is in line with epidemiological estimates (He et al., 2020). We set the model up to return Re for 2 time intervals, as determined from the model. We used Markov Chain Monte Carlo (MCMC) to estimate the parameters of the model with a chain-length of 1 · 10 and sampling every 5000 steps We removed the first 10% of the chain as burnin. We assessed the sampling of the trees using Tracer (v 1.7.1), and made sure that our 2 Re parameters had an effective sample size of at least 100. We ran the model using the genomes from the cluster of B.1.43 strains found in LA County that we inferred arose from a single introduction event collected between the 24th of February to the 19th of April, 2020. This approach is similar to a study of New Zealand SARS-CoV-2 genomes (Geoghegan et al., 2020). For this analysis, we removed a sequence (USA/CA-CSMC31/2020), which caused the initial tree to be unrealistically long and prevented the model from obtaining realistic estimates for when the outbreak started.

